# Submission policy similarity and resubmission burden across the top 50 ophthalmology journals

**DOI:** 10.64898/2026.03.20.26348949

**Authors:** Shayaan Kaleem, Dream Tuitt-Barnes, Oreoluwapo Maxwell, Jonathan A Micieli

## Abstract

After rejection, resubmission of scientific manuscripts often requires substantial journal-specific reformatting. We compared systematic review submission policies across high-impact ophthalmology journals and quantified policy similarity to support resubmission planning. We identified the top 50 ophthalmology journals by SCImago Journal Rank that publish systematic reviews and are not invite-only, extracted policies from author instructions using an a priori data dictionary, and computed pairwise similarity on a 0 to 1 scale using the Gower coefficient across mixed policy variables with available-case denominators for unstated fields. Policies were heterogeneous and frequently unstated. Only 29 of 50 journals (58%) stated a main-text word limit; among journals with numeric limits, the median was 4000 words (interquartile range 3500 to 5500; n = 23). Preferred Reporting Items for Systematic Reviews and Meta-Analyses compliance was explicitly required by 35 of 50 journals (70%), and prospective registration by 6 of 50 journals (12%). Across 1225 journal pairs, similarity was modest, with a median of 0.64 (interquartile range 0.57 to 0.71; range 0.05 to 0.98). Similarity among the top 5 highest-ranking journals ranged from 0.62 to 0.90 (median 0.75). Systematic review submission policies vary widely across high-impact ophthalmology journals, and most journal pairs show only modest similarity. Similarity-based guidance may help identify policy-aligned resubmission targets while anticipating common sources of reformatting burden.

## Introduction

Systematic reviews and meta-analyses are foundational to evidence-based ophthalmic practice, yet submitting them to journals remains operationally complex.[1-3] Beyond scientific rigor, authors must navigate heterogeneous journal requirements for reporting checklists, registration, abstract format, word limits, and submission components.[4] When a systematic review is rejected and resubmitted, these differences can translate into repeated reformatting and administrative effort that is separate from improving scientific content, delaying dissemination and increasing opportunity cost. Empirical studies suggest that even when scientific revisions are minimal, the technical components of resubmission alone typically require several hours per manuscript, with an estimated 3-4 hours per resubmission devoted to reformatting and journal-specific compliance tasks.[5] This avoidable burden consumes substantial researcher time and resources, motivating calls for streamlined or format-free initial submission policies.[6,7] While guidance exists for reporting quality, including Preferred Reporting Items for Systematic reviews and Meta-Analyses (PRISMA) and related extensions, and broader reporting guideline infrastructure such as the Enhancing the QUAlity and Transparency Of health Research (EQUATOR) Network, there is less clarity on how consistently high-impact journals operationalize these expectations within their author guides.[2-4,8] In practice, requirements may be partially specified, presented in general instructions not tailored to systematic reviews, or embedded within broader submission policies. Different journals do not consistently agree on, or enforce, the same reporting standards and this inconsistency makes author instructions vary from journal to journal. This variability is most consequential during resubmission planning, when authors must select a next journal and anticipate the degree of policy mismatch and administrative rework.

We therefore conducted a cross-sectional comparison of systematic review submission requirements across high-impact ophthalmology journals and quantified policy similarity using a mixed-data similarity metric. Our goal was to characterize heterogeneity and incomplete policy specification, quantify similarity within and beyond the top 5 highest impact ophthalmology journals, and provide an evidence-based framework to support resubmission planning.

## Materials and methods

### Journal sampling and eligibility

We began with the SCImago Journal Rank (SJR) portal to identify high-impact ophthalmology journals. SJR is a Scopus-based journal prestige metric that applies an eigenvector-style algorithm to the citation network, such that citations from more influential journals contribute greater weight than citations from less influential journals; the resulting SJR score reflects average journal prestige per article rather than raw citation counts alone.[9] We selected the ophthalmology subject category and screened journals in descending SJR order. We chose SJR because it is field-specific, publicly accessible, and incorporates citation quality (prestige of the citing source) in addition to citation quantity, enabling a transparent and reproducible sampling frame for high-impact journals. Journals were eligible if they explicitly accepted systematic reviews or meta-analyses and did not indicate that systematic reviews were invite-only. Invite-only titles and journals that did not accept systematic reviews were excluded and replaced iteratively by the next highest-ranked eligible journal until 50 eligible journals were identified. The University of Toronto Research Ethics Board ruled that approval was not required for this study.

### Data extraction

Author guides and associated submission instruction pages were reviewed and policies relevant to systematic review submissions were extracted into a predefined spreadsheet on Microsoft Excel with an a priori data dictionary. Extracted fields included reporting and registration (such as PROSPERO) requirements, abstract format and length, manuscript length, and reference limits when stated, limits on figures and tables, preprint policies, required submission components (e.g. cover letter), and submission metadata requirements. Fields not explicitly stated were coded as missing rather than assumed.

### Primary outcome and similarity metrics

The primary outcome was policy similarity between journals. We quantified similarity using the Gower similarity coefficient, which is designed for datasets that contain a mixture of variable types. Gower similarity computes a per-variable similarity on a 0 to 1 scale and then averages these similarities across variables.[10] For categorical and binary variables, similarity was 1 for an exact match and 0 otherwise. For numeric constraints, similarity was calculated as one minus the absolute difference divided by the observed range of that variable across journals, so that larger numeric discrepancies produce lower similarity. We selected Gower similarity because author guide requirements include mixed binary, categorical, and numeric fields, and this approach provides a transparent, interpretable composite similarity score without requiring arbitrary conversion of numeric policies into categories. Overall similarity was computed using available-case denominators, fields missing or unstated for either journal were excluded from the pairwise calculation and the remaining weights were renormalized.

### Anchor journals and resubmission recommendations

We defined “anchor journals” as the top 5 highest SJR journals (Ophthalmology, JAMA Ophthalmology, American Journal of Ophthalmology, Survey of Ophthalmology, and Ophthalmology Retina) and used their submission criteria as a reference to compare other journals against. This was done to simulate how authors often submit to a high impact journal first, and if rejected, reformat for submission to a lower impact journal. For each anchor, candidate journals were ranked by similarity to generate resubmission recommendations.

### Heterogeneity and resubmission friction analyses

We summarized the prevalence of policy fields across journals by reporting medians and interquartile ranges for numeric limits among journals that reported numeric values. To quantify global heterogeneity, we summarized the distribution of similarity across all journal pairs. Practical resubmission friction among policy-aligned targets was quantified by tabulating the most common mismatches observed across the top 10 similarity-based recommendations for each anchor journal.

### Sensitivity analysis

As a secondary analysis, we evaluated whether recommendations were robust to weighting mismatches by anticipated effort. Concordance between similarity-only and effort-weighted rankings was quantified using Spearman correlation and top 5 overlap. All analyses were conducted using Python (version 3.14.2).

## Results

### Study sample and policy completeness

The final dataset included 50 high-impact ophthalmology journals that accept non-invited systematic reviews. Across journals, key submission constraints were frequently missing or specified in non-comparable units, which itself contributes to resubmission uncertainty. For example, 29/50 (58%) journals stated a main-text limit, but only 23 provided a numeric word limit suitable for direct comparison (median 4000; IQR 3500 to 5500; n=23). In contrast, 48/50 (96%) stated an abstract word limit (median 250; IQR 250 to 262.50; n=48). Limits on references, figures, and tables were much less consistently reported, limiting the extent to which authors can anticipate downstream revisions from author guides alone (Table 1).

**Table 1.** Sample characteristics and summary of extracted submission constraints.

| Characteristic | Value, n (%) | Details (when available) |
| --- | --- | --- |
| Main-text word limit stated | 29 (58%) | Median 4000 (IQR 3500-5500; n=23) |
| Abstract word limit stated | 48 (96%) | Median 250 (IQR 250-262.50; n=48) |
| Max references stated | 14 (28%) | Median 70 (IQR 47.50-100; n=12) |
| Max figures stated | 12 (24%) | Median 6 (IQR 5.50-6; n=11) |
| Max tables stated | 10 (20%) | Median 5 (IQR 4-6; n=9) |
| PRISMA explicitly required | 35 (70%) |  |
| Format-flexible initial submission (explicit policy) | 4 (8%) |  |
| Preprints explicitly allowed | 43 (86%) |  |

IQR = Interquartile Range

### Reporting and registration heterogeneity

PRISMA format compliance was explicitly required by 35/50 (70%) journals, whereas registration was explicitly required by 6/50 (12%). Abstract formatting showed additional variability: 37/50 (74%) journals required a structured abstract, while the remainder permitted unstructured formats, allowed either format, or conditioned the structure on article subtype. Preprints were explicitly allowed by 43/50 (86%) journals. Notably, only 4/50 (8%) journals explicitly permitted format-flexible initial submission, suggesting that most resubmissions still require meaningful formatting work even when the science is unchanged (Table 2).

**Table 2.** Format-flexible initial submission policies across journals.

| Journal Ranking | Journal Name | Submission Criteria |
| --- | --- | --- |
| 11 | British Journal of Ophthalmology | Minimal reformatting before editorial review:<br>• Submit an editable manuscript file (e.g., Word) and include abstract, main text with headings, tables, and required statements (e.g. conflicts, funding, ethics).<br>• No journal template or house reference style required at submission; references can be in any consistent format if complete and cited numerically.<br>• If accepted, journal styling is applied during copyediting/proofs. |
| 28 | Ophthalmic and Physiological Optics | Free-format submission (publisher policy):<br>• Submit in the authors' preferred format (no page-layout or house reference style required at initial submission).<br>• Use a consistent reference style with complete citation elements; include required sections, figure/table captions, and disclosures/ethics.<br>• If accepted, the publisher reformats to journal style. |
| 30 | Experimental Eye Research | Format-flexible references at submission:<br>• References may be in any consistent style if complete; journal reference style is applied at proof stage.<br>• Full submission must use editable source files (Word or LaTeX); Word should be single-column; PDF is not an acceptable source file.<br>• Highlights are required as a separate file; tables must be editable text (not images). |
| 37 | BMJ Open Ophthalmology | Minimal reformatting before editorial review:<br>• Submit an editable manuscript file (e.g. Word) and include abstract, main text with headings, tables, and required statements (e.g. conflicts, funding, ethics).<br>• No journal template or house reference style required at submission; references can be in any consistent format if complete and cited numerically. |
|  |  | • If accepted, journal styling is applied during copyediting/proofs. |

### Global similarity and heterogeneity

Across all 1 225 journal pairs, similarity was modest, with a median of 0.64 (IQR 0.57 to 0.71; range 0.05 to 0.98). Most pairs were only moderately aligned: 72% had similarity at or below 0.70 and only 8.5% reached 0.80 or higher.

Similarity among the five anchor journals ranged from 0.62 to 0.90 (median 0.75), indicating partial convergence at the top of the field (Fig 1). However, the typical similarity between an anchor and the broader journal set remained moderate (anchor-specific medians 0.60 to 0.66). Similarity-based rankings identified the most policy-aligned journals for each anchor (Table 3). We also created a dashboard for the highest SJR journal, Ophthalmology, highlighting the specific policy differences authors should anticipate when resubmitting a manuscript to one of the top candidate journals (S1 Table).

**Table 3.**
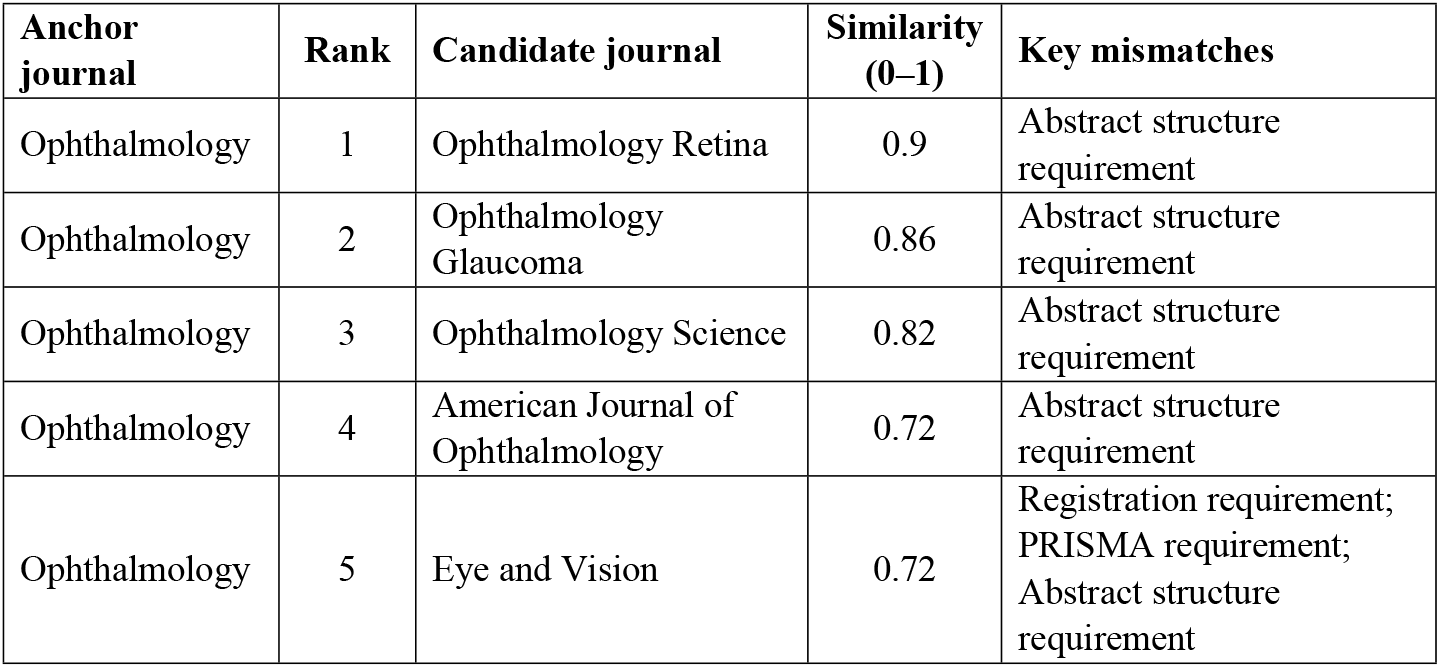
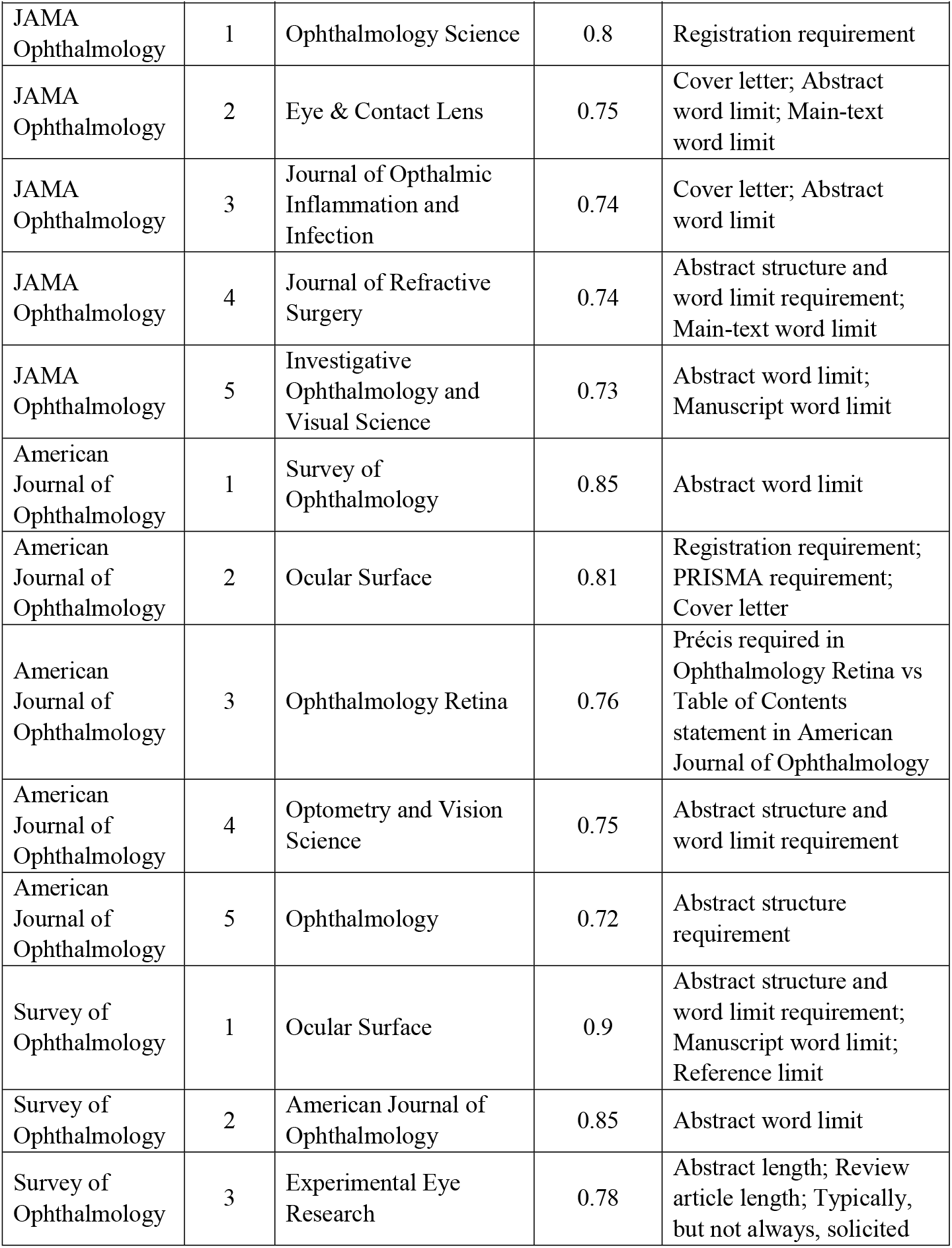
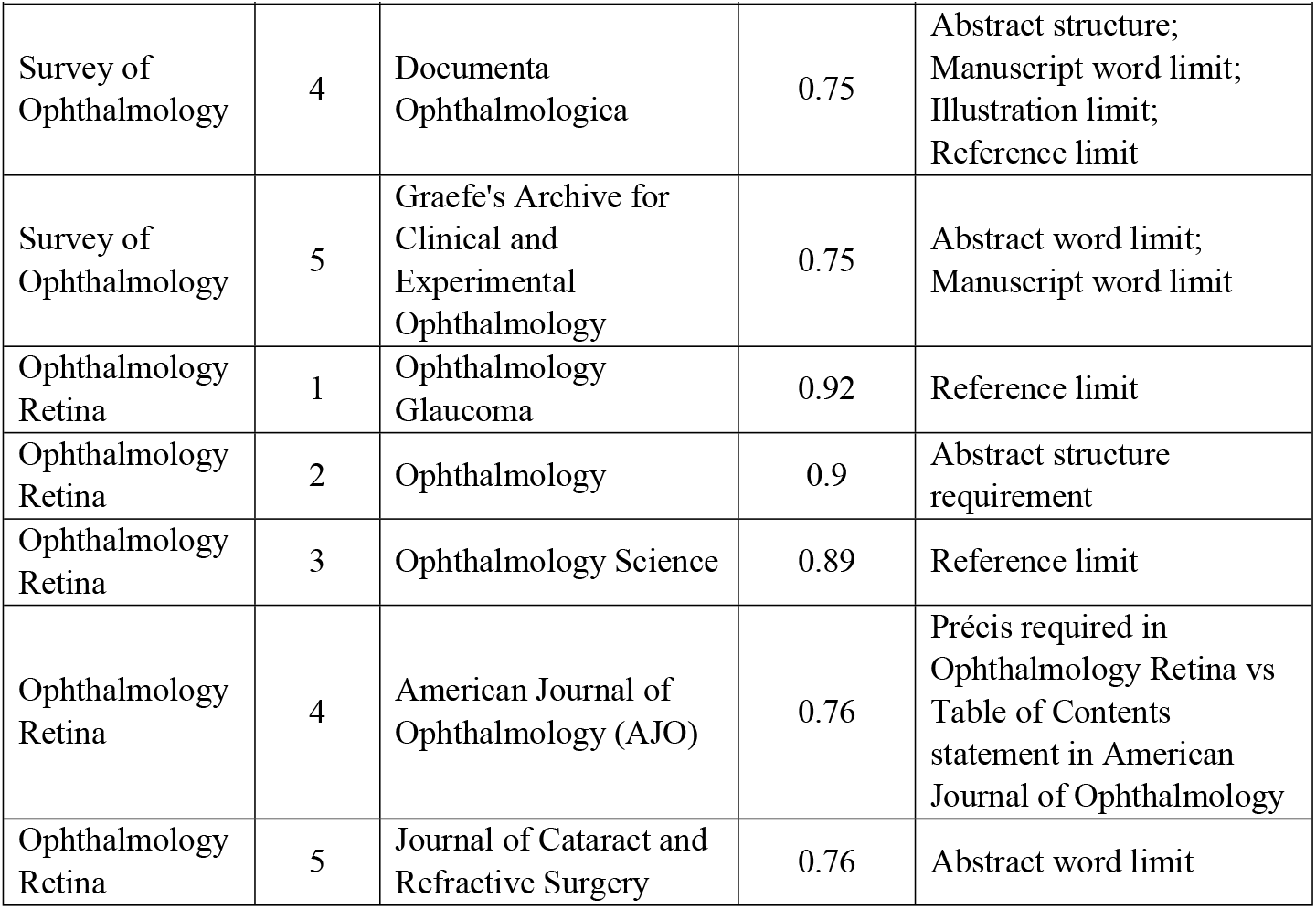
Similarity-based resubmission recommendations for the top 5 highest-impact ophthalmology journals.

| Anchor journal | Rank | Candidate journal | Similarity (0–1) | Key mismatches |
| --- | --- | --- | --- | --- |
| Ophthalmology | 1 | Ophthalmology Retina | 0.9 | Abstract structure requirement |
| Ophthalmology | 2 | Ophthalmology Glaucoma | 0.86 | Abstract structure requirement |
| Ophthalmology | 3 | Ophthalmology Science | 0.82 | Abstract structure requirement |
| Ophthalmology | 4 | American Journal of Ophthalmology | 0.72 | Abstract structure requirement |
| Ophthalmology | 5 | Eye and Vision | 0.72 | Registration requirement; PRISMA requirement; Abstract structure requirement |

| **Anchor journal** | **Rank** | **Candidate journal** | **Similarity (0–1)** | **Key mismatches** |
| --- | --- | --- | --- | --- |
| JAMA Ophthalmology | 1 | Ophthalmology Science | 0.8 | Registration requirement |
| JAMA Ophthalmology | 2 | Eye & Contact Lens | 0.75 | Cover letter; Abstract word limit; Main-text word limit |
| JAMA Ophthalmology | 3 | Journal of Ophthalmic Inflammation and Infection | 0.74 | Cover letter; Abstract word limit |
| JAMA Ophthalmology | 4 | Journal of Refractive Surgery | 0.74 | Abstract structure and word limit requirement; Main-text word limit |
| JAMA Ophthalmology | 5 | Investigative Ophthalmology and Visual Science | 0.73 | Abstract word limit; Manuscript word limit |
| American Journal of Ophthalmology | 1 | Survey of Ophthalmology | 0.85 | Abstract word limit |
| American Journal of Ophthalmology | 2 | Ocular Surface | 0.81 | Registration requirement; PRISMA requirement; Cover letter |
| American Journal of Ophthalmology | 3 | Ophthalmology Retina | 0.76 | Précis required in Ophthalmology Retina vs Table of Contents statement in American Journal of Ophthalmology |
| American Journal of Ophthalmology | 4 | Optometry and Vision Science | 0.75 | Abstract structure and word limit requirement |
| American Journal of Ophthalmology | 5 | Ophthalmology | 0.72 | Abstract structure requirement |
| Survey of Ophthalmology | 1 | Ocular Surface | 0.9 | Abstract structure and word limit requirement; Manuscript word limit; Reference limit |
| Survey of Ophthalmology | 2 | American Journal of Ophthalmology | 0.85 | Abstract word limit |
| Survey of Ophthalmology | 3 | Experimental Eye Research | 0.78 | Abstract length; Review article length; Typically, but not always, solicited |
| Survey of Ophthalmology | 4 | Documenta Ophthalmologica | 0.75 | Abstract structure;<br>Manuscript word limit;<br>Illustration limit;<br>Reference limit |
| Survey of Ophthalmology | 5 | Graefe's Archive for Clinical and Experimental Ophthalmology | 0.75 | Abstract word limit;<br>Manuscript word limit |
| Ophthalmology Retina | 1 | Ophthalmology Glaucoma | 0.92 | Reference limit |
| Ophthalmology Retina | 2 | Ophthalmology | 0.9 | Abstract structure requirement |
| Ophthalmology Retina | 3 | Ophthalmology Science | 0.89 | Reference limit |
| Ophthalmology Retina | 4 | American Journal of Ophthalmology (AJO) | 0.76 | Précis required in Ophthalmology Retina vs Table of Contents statement in American Journal of Ophthalmology |
| Ophthalmology Retina | 5 | Journal of Cataract and Refractive Surgery | 0.76 | Abstract word limit |

**Fig 1.**
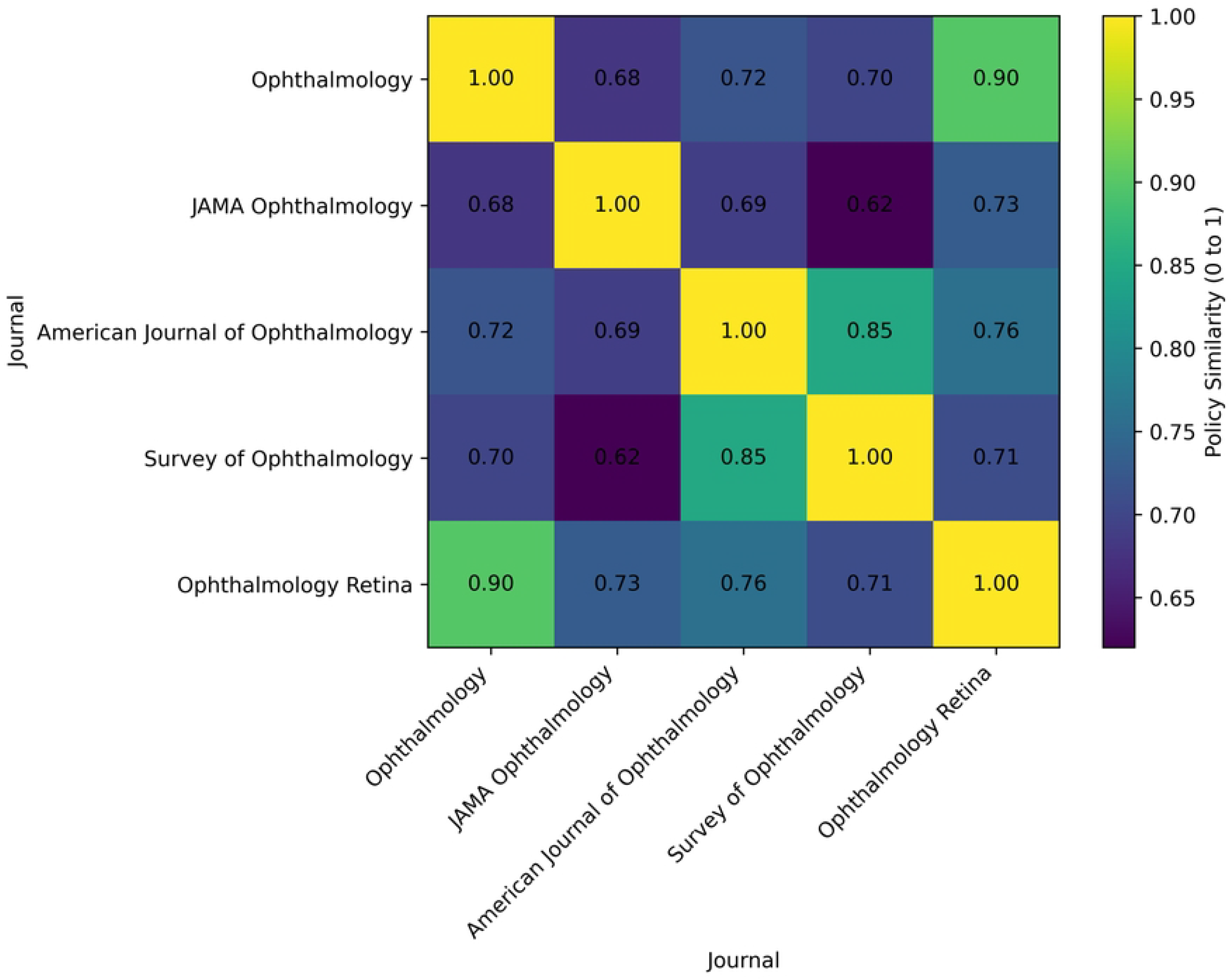
Policy similarity heatmap among the top 5 highest-impact ophthalmology journals. Residual resubmission friction among policy-aligned targets.

Even among top similarity-based recommendations, mismatches were common (S1 Fig). Across the top 10 candidate journals for each anchor (n=50 anchor-target pairs), the most frequent mismatches involved abstract word limits (40%), registration requirements (30%), and abstract structure (26%), followed by cover letter requirements (12%). These mismatches represent predictable sources of reformatting and additional documentation at resubmission, even when selecting a policy-similar target.

### Sensitivity analysis

In sensitivity analyses that weighted policy mismatches by anticipated effort, similarity-only recommendations were broadly consistent with effort-weighted rankings when pooled across both weighting schemes (mild and strong) using the lower concordance for each anchor (Spearman rho 0.80 to 0.96; Fig 2). The number of journals that were top 5 in similarity to each anchor journal before and after weighting were: 2/5 for Ophthalmology and JAMA Ophthalmology, 4/5 for American Journal of Ophthalmology and Ophthalmology Retina, and 5/5 for Survey of Ophthalmology. When examined separately, the mild weighting yielded Spearman rho 0.80 to 0.96 with a top 5 overlap of 2/5 to 5/5, whereas the strong scheme yielded Spearman rho 0.89 to 1.00 with a top 5 overlap of 4/5 to 5/5. The weighting for each policy across the three weighting methods (unweighted, mild, and strong) can be found in S2 Table.

**Fig 2.**
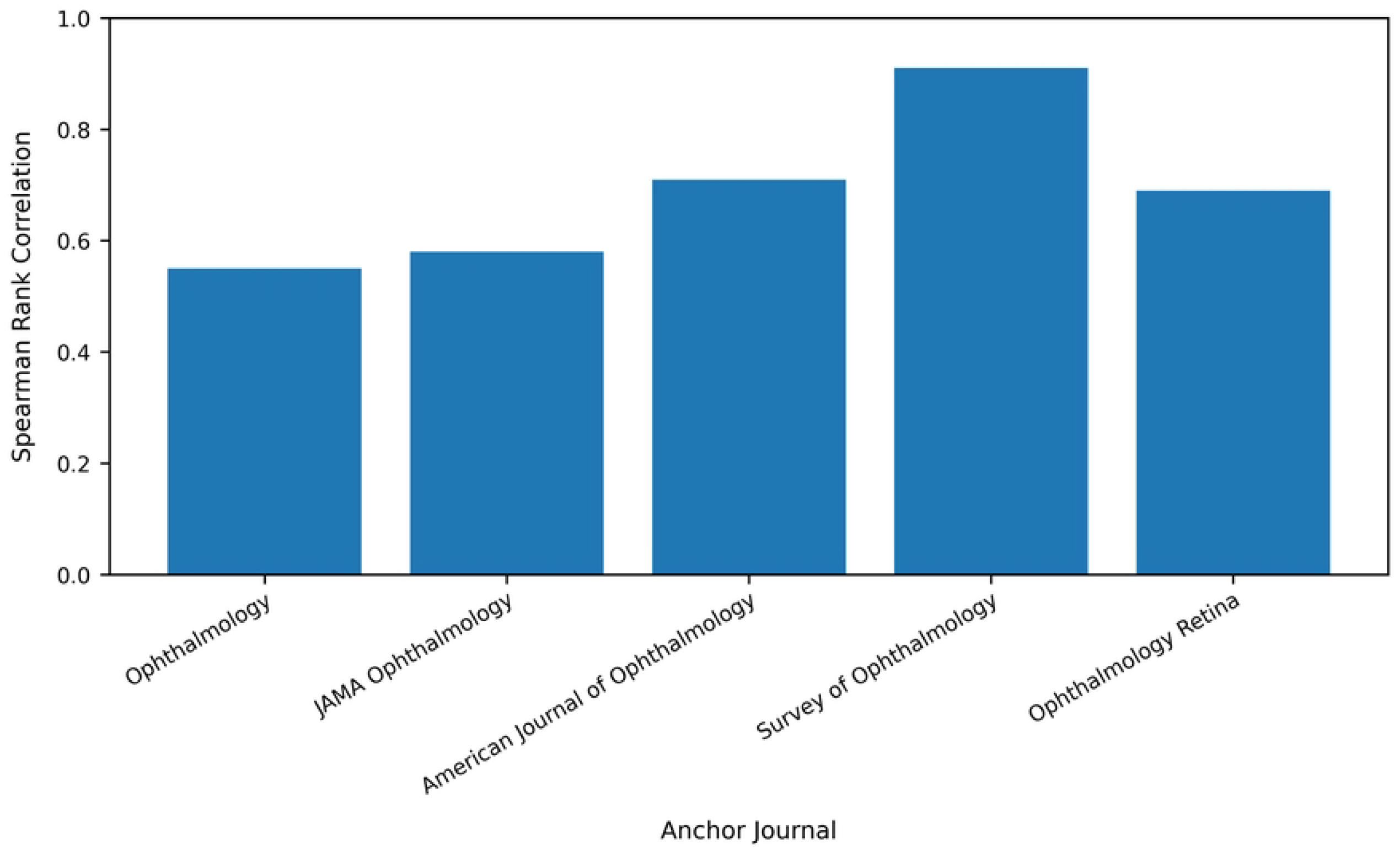
Spearman correlation comparing the top 5 ophthalmology journal similarity to all other journals with and without effort weighting.

## Discussion

In this comparative analysis of the top 50 high-impact ophthalmology journals that publish non-invited systematic reviews, we found substantial heterogeneity in author guide requirements and frequently incomplete specification of key policies. This heterogeneity is not a minor formatting nuance: most journal pairs showed only modest similarity and only a small fraction were highly aligned. In the broader biomedical literature, variation in submission requirements within the same scientific focus has been shown to impose a sizable resubmission burden, including measurable delays to publication.[11] Moreover, the recurring mismatches in abstract limits, registration requirements, and abstract structure among policy-aligned resubmission targets found in our study indicate that resubmission effort often remains meaningful.

Beyond conceptual inefficiency, resubmission following rejection imposes a measurable time and labor burden on authors. Survey data indicates that the technical components of resubmission, including manuscript reformatting and compliance with journal-specific requirements, commonly requires several hours to multiple days per resubmission, with approximately 65% of authors reporting one to three days or more spent on resubmission logistics after rejection.[6] While resubmission effort may also include revisions in response to editorial or peer-review feedback, prior work suggests that formatting and policy compliance alone contribute substantially to resubmission delays, with reported delays exceeding two weeks for most manuscripts and over three months for approximately 20%.[6] When aggregated across the volume of rejected manuscripts, these burdens translate to millions of researcher-hours annually and an estimated economic cost exceeding $1 billion United States dollars per year.[6] Another study estimated that manuscript reformatting consumes a median of 52 hours per researcher per year.[12]

These findings have practical implications for authors and research groups. Author guides frequently omit or incompletely specify constraints that drive major edits, such as reference caps or figure limits, which can amplify uncertainty and downstream rework. When constraints are specified, they are not always stated in comparable units (for example pages versus words), which complicates planning and may necessitate additional author-editor clarification. Explicit format-flexible submission policies were uncommon, suggesting that the default expectation remains full compliance with journal-specific formatting at initial submission, even though many editorial systems can accommodate post-acceptance standardization.[6,7] Together, these features create predictable friction during resubmission, particularly for time-sensitive evidence syntheses.

Our similarity framework helps operationalize this problem. Rather than relying on informal heuristics, authors can quantify how closely a candidate journal matches the requirements of the source journal and can anticipate the most likely sources of mismatch using field-level summaries. The sensitivity analysis suggests that recommendations are not highly sensitive to whether mismatches are treated as equally important or weighted by anticipated effort, supporting the defensibility of this similarity-based analysis as a baseline resubmission strategy. From an editorial and publisher perspective, our results support actionable opportunities to reduce avoidable author burden without compromising reporting rigor. Clear, review-specific author guide sections that explicitly state requirements for PRISMA compliance, registration expectations, abstract structure, and key limits would improve transparency and may improve adherence to established reporting standards.[2,3,8] Wider adoption of format-flexible initial submission policies could reduce administrative workload for both authors and editorial offices, concentrating effort on scientific merit and reporting completeness.[6,7] Harmonizing a small set of systematic review submission elements across journals could meaningfully reduce friction while preserving journal identity and editorial standards, aligning with broader efforts to standardize core expectations in biomedical publishing.[4]

Heterogeneity in submission policies may also create inequitable administrative burden by increasing reformatting and compliance work during resubmission. This workload may disproportionately affect early career researchers, trainees, clinician scientists with limited protected time, and authors without dedicated editorial support, including in low- and middle-income settings. Similarity based resubmission planning may improve accessibility by making policy differences more transparent and helping authors prioritize scientifically substantive revision over avoidable administrative compliance.

This study has notable limitations. First, author guide requirements can change over time, and our extraction reflects a cross-sectional snapshot. Second, policies were coded from publicly available instructions and may not fully capture editorial discretion, submission system prompts, or case-by-case guidance provided during peer review. Third, missingness in author guides may underestimate true heterogeneity if policies exist but are not documented, as our available-case approach avoids imputing unstated requirements. Finally, time spent on resubmission was not directly measured; hence we were unable to quantify if greater dissimilarity between journal policies represents a significantly greater time spent on resubmission. Although we aimed to weight submission policies by the amount of effort they would require to complete, individual manuscript and author differences may make certain policies more time consuming to certain individuals or articles than to others.

Overall, systematic review submission policies differ substantially across high-impact ophthalmology journals and most pairs of journals exhibit only modest policy similarity. This heterogeneity and incomplete policy specification are likely to generate avoidable resubmission effort. This similarity-based resubmission framework can guide selection of policy-aligned journals for resubmission while making expected mismatches transparent to authors.

## Data Availability

All data underlying the findings of this study are derived from publicly available journal submission guidelines and author instructions available on the official websites of the respective journals. Because these sources are publicly accessible and the dataset represents a compiled extraction of this information, the data can be independently reconstructed. The extracted dataset used in the analyses is available from the corresponding author upon reasonable request.

## Acknowledgements

None.

## Supporting information captions

**S1 Table. Ophthalmology-specific dashboard for the highest-ranked similarity-based targets**. ^a^ Requirement differs from Ophthalmology

**S2 Table. Policy weights used across the unweighted, mild, and strong weighting schemes**. ^a^ Weights were applied to the primary Gower similarity calculation to reflect the anticipated relative effort required to comply with each policy after rejection. Unweighted analysis assigned weight 1.00 to all variables.

**S1 Fig. Most frequent mismatches across the top 10 candidate journals for each anchor journal**.

## Notes

### Competing Interest Statement

The authors have declared no competing interest.

### Funding Statement

The author(s) received no specific funding for this work.

## References

1. Li T, Bartley GB. Publishing systematic reviews in ophthalmology: new guidance for authors. Ophthalmology. 2014;121:438–9.

2. Page MJ, McKenzie JE, Bossuyt PM, et al. The PRISMA 2020 statement: an updated guideline for reporting systematic reviews. BMJ. 2021;372:n71.

3. Simera I, Moher D, Hirst A, et al. Transparent and accurate reporting increases reliability, utility, and impact of your research: reporting guidelines and the EQUATOR Network. BMC Med. 2010;8:24.

4. Hopewell S, Altman DG, Moher D, Schulz KF. Endorsement of the CONSORT statement by high impact factor medical journals: a survey of journal editors and journal ‘Instructions to Authors’. Trials. 2008;9:20.

5. Sobani ZA, Horovitz J, Kamholz S. Streamlined manuscript submission guidelines: beyond overdue. Ann Med Surg (Lond). 2018;25:1–2.

6. Jiang Y, Lerrigo R, Ullah A, et al. The high resource impact of reformatting requirements for scientific papers. PLoS One. 2019;14:e0223976.

7. Clotworthy A, Davies M, Cadman TJ, et al. Saving time and money in biomedical publishing: the case for free-format submissions with minimal requirements. BMC Med. 2023;21:172.

8. Moher D, Liberati A, Tetzlaff J, Altman DG; PRISMA Group. Preferred reporting items for systematic reviews and meta-analyses: the PRISMA statement. PLoS Med. 2009;6:e1000097.

9. González-Pereira B, Guerrero-Bote VP, Moya-Anegón F. A new approach to the metric of journals’ scientific prestige: the SJR indicator. J Informetr. 2010;4:379–91.

10. Gower JC. A general coefficient of similarity and some of its properties. Biometrics. 1971;27:857–71.

11. Malički M, Aalbersberg IJJ, Bouter LM, ter Riet G. Journals’ instructions to authors: a cross-sectional study across scientific disciplines. PLoS One. 2019;14:e0222157.

12. LeBlanc AG, Barnes JD, Saunders TJ, Tremblay MS, Chaput JP. Scientific sinkhole: the pernicious price of formatting. PLoS One. 2019;14:e0223116.

